# From Health Needs to the Use of Health Services: A Scoping Review Protocol on Access to Medicines

**DOI:** 10.1101/2025.10.29.25339098

**Authors:** Helena Luiza Kirsten Sasse, Maria Eduarda Pereira, Mônica Cristina Nunes, Ernani Tiaraju de Santa Helena, Luana Gabriele Nilson

**Author notes:** **CORRESPONDING AUTHOR:** Helena Luiza Kirsten Sasse.

## Abstract

This study aims to analyze the relationship between health needs (felt and expressed) and access to medicines. It is a scoping review based on the Joanna Briggs Institute (2024) guidelines, structured according to the PCC strategy (Population, Concept, and Context). Searches will be conducted in PubMed, SciELO, LILACS, Scopus, and Embase for articles published between 2015 and 2025, in Portuguese, English, Spanish, and German. Inclusion criteria comprise primary studies and systematic reviews addressing health needs in relation to access to medicines, while clinical trials, editorials, letters, and commentaries will be excluded. Search keys were developed according to the classification of types of needs. Study selection will be carried out by two independent reviewers using the Rayyan© platform, and results will be presented through the PRISMA-ScR flowchart. In cases of disagreement, a third reviewer will evaluate the conflicts. Data extracted from the included studies will be organized in a chart containing information such as title, country, year, authors, journal, study design, objectives, and main findings.

## 1. INTRODUCTION

The use of health services is central to the functioning of health systems, as the utilization process results from the interaction between the behavior of individuals seeking care and the professionals guiding them within the system (Poças et al., 2019).

Access to health services can be described as a set of dimensions that mediate the relationship between demand and entry into the service, constituting one of the main factors for assessing the quality and performance of health systems (Almeida et al., 2017). Assessing socioeconomic inequalities over the years in terms of access to and quality of health services is an important strategy for promoting equitable access to health services (Coube et al., 2023).

Geographical, financial, organizational, informational, and cultural barriers, among others, reflect supply-side characteristics that facilitate or hinder people’s ability to use health services (Gong et al., 2016; Gliedt et al., 2023). In many countries, including highly developed ones, inequalities in access persist, and studies show that the use of health systems can vary considerably according to individual and system characteristics, such as age, gender, income, race/skin color, educational level, type of coverage, and type of service used (Tille et al., 2017; Abera Abaerei et al., 2017; Gong et al., 2016).

Bradshaw (1972) categorizes social needs, including health needs, into four types: normative (defined by professional standards), felt (related to individual desire), expressed (also referred to as demand, when desire is transformed into action), and comparative (deficits observed in one population group compared with others with similar characteristics). Bradshaw’s taxonomy of social need is useful for understanding different assessments of the value of medicines (Vargas-Peláez et al., 2017).

In this context, access to medicines becomes an indicator of the quality and responsiveness of the health system, with its absence interpreted as a weakening of the processes of cure, rehabilitation, and disease prevention (Boing et al., 2022).

### 1.1 Object of study

This study aims to analyze the relationship between the determinants of health service utilization from the perspective of access to medicines.

## 2. METHODS

### 2.1 Type, period, and study setting

This is a Scoping Review, based on the guidelines of the *Joanna Briggs Institute Reviewers’ Manual* (Aromataris et al., 2024). The review will follow the following steps: To construct the guiding question, the PCC strategy (Population, Concept, Context) was used, being P: Population – Individuals who use health services; C: Concept – Health needs (felt and expressed) and their relationship with the utilization of health services; C: Context – Access to medicines. Thus, the following research question was defined: Is there a relationship between the different health needs (felt and expressed) and access to medicines?

### 2.2 Eligibility criteria

The inclusion and exclusion criteria are described in Table 1, with emphasis on the thematic focus on health needs (felt and expressed) and their relationship with access to medicines (with needs considered according to the concepts previously presented and represented by the search keys).

**Table 1:** Selection Criteria.

| Selection Criteria | Inclusion Criteria | Exclusion Criteria |
| --- | --- | --- |
| Period | 2015 a 2025 | Anteriores a 2015 |
| Language | Portuguese, English, Spanish, German | Other languages |
| Study Type | Primary studies (qualitative and quantitative), Systematic reviews | Articles not available in full text; duplicates; clinical trials; editorials; reviews that are not systematic; letters to the editor; commentaries; research; protocols. |
| **Topics** | Health needs (felt and expressed); access to medicines; pharmaceutical services; use of health services | Studies that do not address health needs or access to medicines; studies that only focus on clinical efficacy of medicines without discussing access; studies that address only the use of health services or health needs (felt and expressed) without discussing access to medicines. |

Source: Authors, 2025.

### 2.3 Search strategy

Searches will be conducted through the following databases: PubMed, SciELO, LILACS, Scopus, and Embase.

For conducting the research, the classification of needs will follow the theoretical model of Vargas-Peláez et al. (2017), to compose the following search strategy, described in detail in Table 2:

**Table 2:**
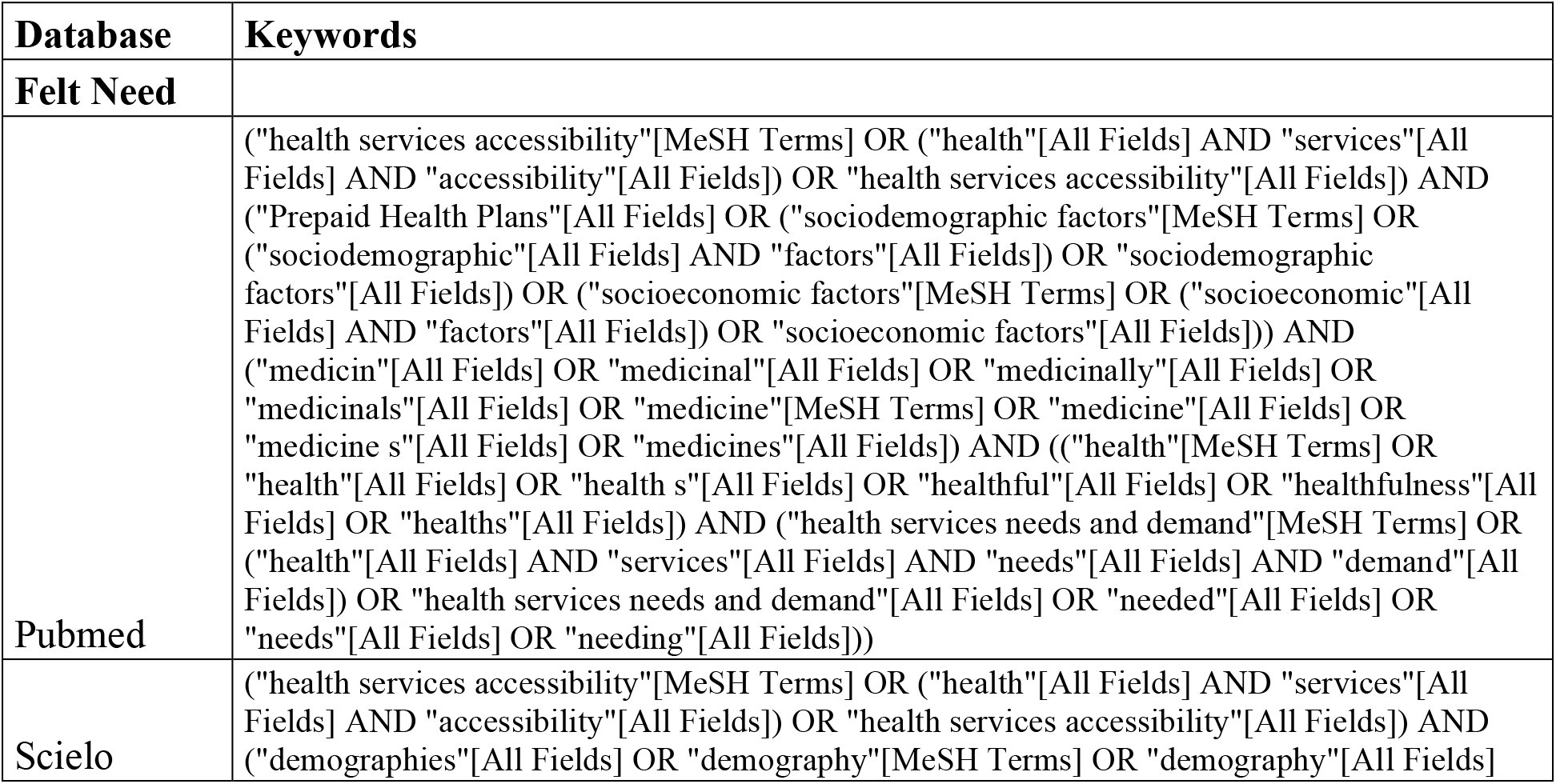
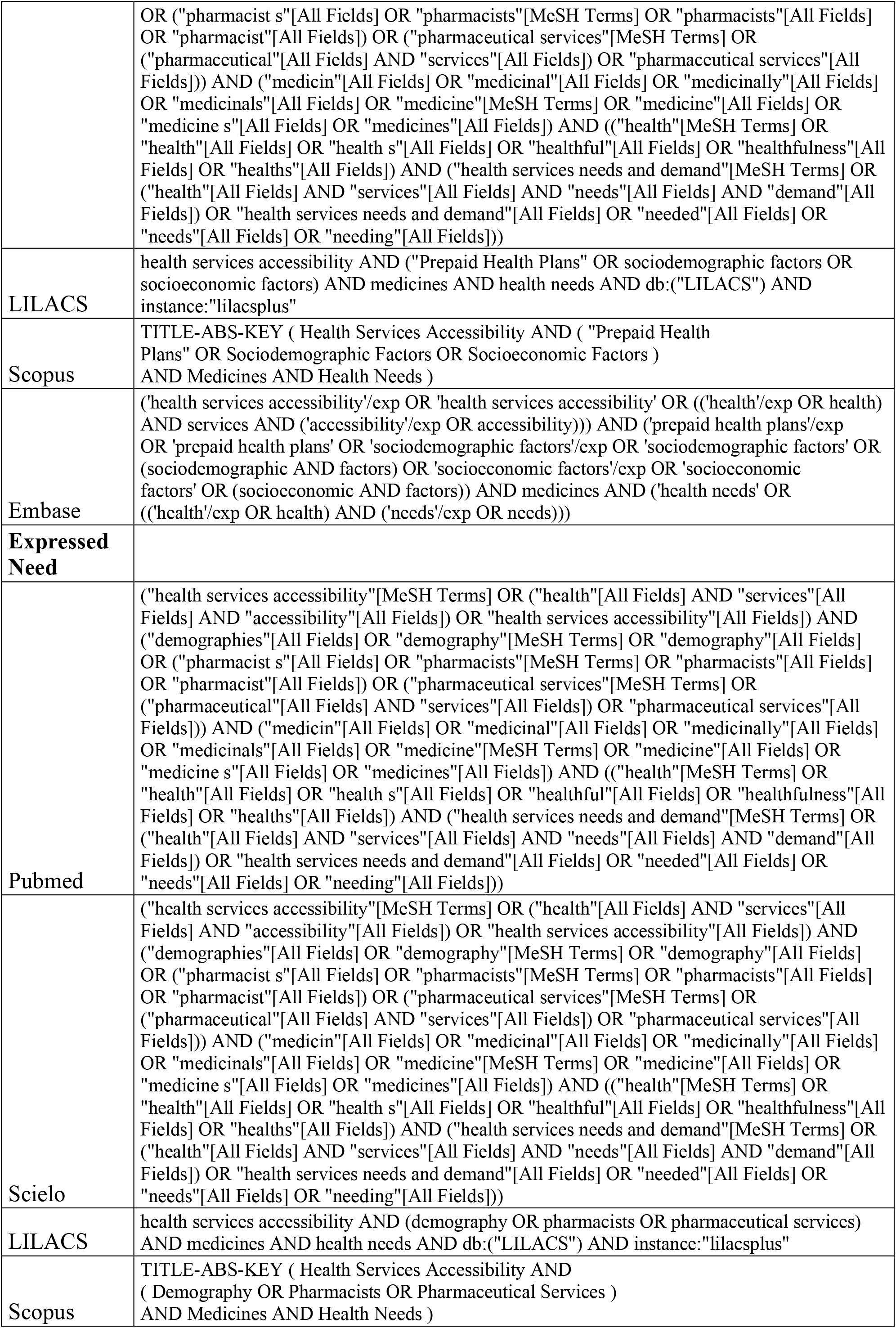
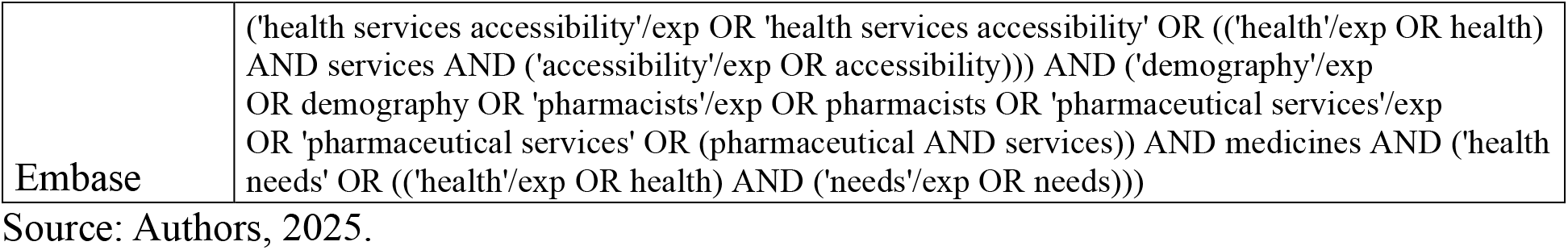
Search Strategy by Database.

#### 1. Felt Need

Health Services Accessibility AND (“Prepaid Health Plans” OR Sociodemographic Factors OR Socioeconomic Factors) AND Medicines AND Health Needs.

#### 2. Expressed Need

Health Services Accessibility AND (Demography OR Pharmacists OR Pharmaceutical Services) AND Medicines AND Health Needs.

In addition to the search keys, filters related to period and language will also be applied.

### 2.4 Study selection

The studies retrieved from the databases will be grouped and uploaded to the Rayyan© platform, also aiming to remove duplicates. After that, the studies will be screened by title and abstract by two independent reviewers according to the inclusion and exclusion criteria. Articles selected in the previous step will be retrieved in full text and evaluated by two reviewers. Articles that are not available in full text or that do not meet the inclusion and exclusion criteria will be excluded, and the process will be presented in a PRISMA flowchart. In cases of disagreement among reviewers, a third reviewer will be consulted for the final decision.

### 2.5 Data analysis and presentation

The data extracted from the articles by the two reviewers will be mapped, including specific details such as: author(s), year, type of publication, country of origin, study design, objectives, and results. This process will ensure that all relevant findings are captured to compose the scoping review. Discrepancies between the two reviewers will be discussed together with a third reviewer.

The results will be presented according to a PRISMA-ScR flowchart, including the inclusion and exclusion criteria. A table will be presented with the following information: study title, country, year, authors, journal, study design, objectives, and results.

### 2.6 Ethical aspects

As it is a scoping review, the project does not need to be submitted to the Research Ethics Committee, but it will follow all the rigor for preparing the research protocol.

## Data Availability

This study did not generate any new data. All data analyzed are from previously published studies.

